# Identifying high-risk pre-term pregnancies using the fetal heart rate and machine learning

**DOI:** 10.1101/2024.02.26.24303280

**Authors:** Gabriel Davis Jones, William Cooke, Manu Vatish

**Affiliations:** Nuffield Department of Women’s & Reproductive Health, University of Oxford; The Alan Turing Institute, London

## Abstract

**Introduction:** Fetal heart rate (FHR) monitoring is one of the commonest and most affordable tests performed during pregnancy worldwide. It is critical for evaluating the health status of the baby, providing real-time insights into the physiology of the fetus. While the relationship between patterns in these signals and adverse pregnancy outcomes is well-established, human identification of these complex patterns remains sub-optimal, with experts often failing to recognise babies at high-risk of outcomes such as asphyxia, growth restriction and stillbirth. These outcomes are especially relevant in low- and middle-income countries where an estimated 98% of perinatal deaths occur. Pre-term birth complications are also the leading cause of death in children ¡5 years of age, 75% of which can be prevented. While advances have been made in developing low-cost digital solutions for antenatal fetal monitoring, there is still substantial progress to be made in developing tools for the identification of high-risk, adverse outcome pre-term pregnancies using these FHR systems. In this study, we have developed the first machine learning algorithm for the identification of high-risk pre-term pregnancies with associated adverse outcomes using fetal heart rate monitoring.

**Methods:** We sourced antepartum fetal heart rate traces from high-risk, preterm pregnancies that were assigned at least one of ten adverse conditions. These were matched with normal pregnancies delivered at term. Using an automated, clinically-validated algorithm, seven distinct fetal heart rate patterns were extracted from each trace, subsequently filtered for outliers and normalized. The data were split into 80% for model development and 20% for validation. Six machine learning algorithms were trained using k-fold cross-validation to identify each trace as either normal or high-risk preterm. The best-performing algorithm was further evaluated using the validation dataset based on metrics including the AUC, sensitivity, and specificity at three distinct classification thresholds. Additional assessments included decision curve analysis and gestational age-specific and outcome-specific performance evaluations.

**Results:** We analysed antepartum fetal heart rate recordings from 4,867 high-risk, pre-term pregnancies with adverse outcomes and 4,014 normal pregnancies. Feature extraction and preprocessing revealed significant differences between the groups (p<0.001). The random forest classifier was the most effective model, achieving an AUC of 0.88 (95% CI 0.87–0.88). When evaluating specific adverse outcomes, the median AUC was 0.85 (IQR 0.81–0.89) and the model consistently exceeded an AUC of 0.80 across all gestational ages. The model’s robustness was confirmed on the validation dataset with an AUC of 0.88 (95% CI 0.86–0.90) and a Brier score of 0.14. Decision curve analysis showed the model surpassed both the treat-none and treat-all strategies over most probability thresholds (0.11–1.0). Performance metrics when using the Youden index were as follows: sensitivity 76.2% (95% CI 72.6–80.5%), specificity 87.5% (95% CI 83.3–91.0), F1 score 81.7 (95% CI 79.6–83.9), and Cohen’s kappa 62.8 (95% CI 59.6–66.4), indicating high discriminative ability between pregnancy outcomes.

**Conclusions:** Our study successfully demonstrated machine learning algorithms are capable of identifying high-risk preterm pregnancies with associated adverse outcomes through fetal heart rate monitoring. These findings demonstrate the potential of machine learning in enhancing the accuracy and effectiveness of antenatal fetal monitoring, particularly for high-risk cases where timely intervention is crucial. This algorithm could substantially improve pregnancy outcome prediction and consequently, maternal and neonatal care, especially in low-to middle-income countries where the burden of adverse outcomes is high.

## Introduction

Monitoring of the fetal heart rate is one of the commonest obstetric investigations worldwide, estimated to be used in more than 85% of pregnancies.[1] Fetal heart rate monitoring (“non-stress test” or “cardiotocography”) involves the non-invasive application of an ultrasound transducer to the maternal abdomen to continuously evaluate the fetal heart rate, enabling real-time, continuous assessment of fetal physiology. Patterns within the fetal heart rate are associated with central and peripheral nervous system and fetal endocrine activity.[2, 3] These patterns are therefore used to evaluate fetal brain health and overall wellbeing.[4, 5] Monitoring in the third trimester before labour (the antepartum) is frequently performed to assess whether a baby is at risk of an adverse outcome or death, indicating early pregnancy intervention is required.[6–8]

Antepartum fetal heart rate monitoring has been a mainstay in pregnancy care since the 1960s, however the reproducibility and reliability of human visual analysis has remained consistently poor.[9] Studies demonstrate expert clinical evaluation fails to accurately identify between 35–92% of fetal heart rate patterns.[10, 11] Inter- and intra-observer agreement between experts has also been estimated as low as 29% while false positive rates for identifying an at-risk fetus are as high as 60%.[12–17] Human misinterpretation of these patterns has therefore been associated with avoidable early pregnancy intervention, increased adverse pregnancy outcomes (including fetal death) and is a major source of medicolegal litigation globally.[18–21] Efforts to standardize visual evaluation methods in antepartum fetal heart rate monitoring have faced issues with performance, reproducibility and clinician consensus. [21–25]

Globally, It is estimated that 98% of perinatal deaths occur in low- and middle-income countries.[26] While technologies for the assessment of fetal wellbeing (e.g. fetal electrocardiography, ultrasound imaging and fetal magnetocardiography) are often prohibitively expensive, fetal heart rate monitoring is one of the most affordable solutions.[26, 27] Recent efforts have also resulted in the development of significantly cheaper mobile-based technologies.[28] The technical training required for practitioners to use these devices is low. However, training users to evaluate these complex signals is frequently a costly barrier to implementation.

Developments in applied machine learning and the curation of large clinical datasets have demonstrated considerable potential for the early detection of pregnancy disorders.[29–31] Machine learning models can now out-perform clinical experts at diagnosis and imaging analysis.[32–34] While machine learning studies have been undertaken for fetal heart rate analysis during labour and delivery (where these patterns vary significantly to the antepartum)[35–37], there is a paucity of such exploration in antepartum, pre-term (<37 weeks) pregnancies. This period in pregnancy is recognised as an area of high need for investigation.[38] Pre-term birth complications are the leading cause of death in children <5 years of age, accounting for an estimated 1 million global deaths annually, with 75% being preventable.[39, 40] The successful application of machine learning to one of the commonest and accessible investigations in pregnancy could contribute substantially to alleviating this burden. It is not yet known if the application of machine learning to antepartum fetal heart rate patterns can identify high-risk pre-term pregnancies. In this study, we have developed a large cohort of high-risk pre-term pregnancies and demonstrated that machine learning algorithms trained on established fetal heart rate patterns possess substantial potential in identifying high-risk pregnancies.

## Methods

### Data processing, study group identification and extraction of fetal heart rate features

We extracted raw digital antepartum fetal heart rate traces from the Oxford University Hospitals maternity database at the John Radcliffe Hospital (Oxford, United Kingdom) between the 30^th^ of November 1990 and 31^st^ of December 2021. Traces were acquired from singleton pregnancies between 27^+0^ and 36^+6^ gestational weeks for which associated clinical outcome information for the mother and baby were available. We then developed a normal cohort of healthy pregnancies delivered at term and a high-risk adverse outcome cohort of pre-term delivery pregnancies. Inclusion and exclusion criteria were used to obtain a normal cohort (Supplementary Table 1). These included records from pregnant women aged between 18–39 years with a BMI ≤30 kg/m^2^, normal pregnancy biomarker and ultrasound scan results, term delivery (37^+0^–41^+0^ weeks), birthweights between 25^th^–75^th^ centiles, normal Apgar scores (≥4 at 1 minute; ≥7 at 5 minutes) and no requirement for neonatal resuscitation or special care admission following delivery. For pregnancies with more than one trace available in a gestational week, only the first trace was used to remove potential bias introduced by subsequent traces that may have been performed due to observations from the initial trace. The high-risk preterm adverse outcome cohort comprised pregnancies in which the baby was classified at delivery as having at least one of low acidaemia, antepartum/intrapartum stillbirth, asphyxia, a birthweight ≤3^rd^ centile for gestational age[41], an extended special care admission ≥7 days, hypoxaemic ischaemic encephalopathy, low Apgar scores, neonatal sepsis, perinatal infections or respiratory conditions. The definition of acidaemia was an arterial pH <7.13 and arterial base deficit >10.0 for babies delivered via caesarean section without labour or arterial pH <7.05 and arterial base deficit >14.0 for babies who experienced labour (regardless of delivery method) in accordance with hospital guidelines where the data were acquired. Low Apgar scores were defined as <4 at 1 minute and <7 at 5 minutes.[42] Asphyxia was defined as low Apgar scores in the presence of acidaemia. Hypoxaemic ischaemic encephalopathy and neonatal sepsis were diagnosed by board-certified neonatologists. Diagnoses were obtained either directly from clinical records or using Phecodes (Phecode version 1.2; perinatal infection: 657, respiratory conditions: 656.2).[43]

We excluded records from babies delivered with inadequate outcome information to avoid potential confounding. FHR monitoring is used to inform clinical decision making, sometimes resulting in avoidable early delivery of the baby. As such, introducing traces from pre-term deliveries in the absence of other independently verifiable adverse outcomes could introduce bias. We then excluded from the high-risk preterm adverse outcome cohort traces that were acquired more than 7 days prior to delivery. Traces are acquired for myriad indications throughout pregnancy. It is therefore unreliable to assume traces acquired throughout pregnancy are for a consistent indication. Incorporating traces acquired substantially earlier than the outcome was identified without clinical evidence would assume all traces acquired for that pregnancy were performed while pathology was present in the fetus. Constraining the time window for adverse outcome traces to within 7 days prior to delivery assists in avoiding this assumption.

We processed the raw antepartum FHR signals with an established automated feature identification algorithm to extract seven features[34, 35]: basal fetal heart rate, accelerations, decelerations, most lost beats (MLB), short-term variation (STV), time spent in an episode of high variation and time spent in an episode of low variation (minutes). Features were not extracted beyond 60 minutes of the trace. These features and their extraction methods have previously been described and clinically validated in the literature.[44] We provide here a brief description of these features. The first procedure in FHR feature extraction is fitting a baseline (the average heart rate excluding any major deviations) to the signal. This serves as the reference point for the trace, facilitating the identification of other features. Accelerations and decelerations are transient deviations above or below this baseline. An acceleration is defined as a temporary increase in the FHR at least 10 bpm above the baseline lasting longer than 15 seconds. A deceleration is a decrease of at least 20 bpm lasting longer than 30 seconds or at least 10 bpm lasting longer than 60 seconds. “Lost beats” is the product of the duration of the deceleration and the magnitude of the deviation from the baseline of deceleration. “Most lost beats” is the largest observed loss of beats due to a deceleration in a FHR trace. STV is the mean absolute difference in time intervals between successive heart rate pulses. Episodes of high and low variation are defined as episodes in which the variability of the FHR trace is consistently above (high variation) or below (low variation) pre-determined thresholds of variation. Fetal movements were not included in our analysis, as they are a subjective measurement and often not recorded in a clinical setting. We then examined each trace for outliers, excluding any trace that demonstrated >30% signal loss, basal fetal heart rate <100 or >180bpm, >1 acceleration per minute, >125 most lost beats, or an STV <2 or >30 milliseconds based on clinical expertise. Each FHR trace feature was then transformed into either a z-score (where the distribution approximated normal) or using min-max normalisation. We performed propensity score matching, matching for gestational age at FHR trace acquisition, fetal sex and trace duration. The K-nearest neighbours algorithm was used to sample without replacement, matching each identified case of a preterm adverse outcome to a normal outcome pregnancy where available. The data were then randomly spliced 80:20% into a model training dataset and internal validation dataset, balanced for outcome, gestational age, trace duration and fetal sex.

### Development of machine learning models

We trained six machine learning algorithms on the model training dataset to predict whether a trace belonged to the normal or preterm adverse outcome pregnancy. The algorithms were a decision tree (DT), Gaussian naïve Bayes (GNB), logistic regression (LR), random forest (RF), support vector machine (SVM) and XGBoost (XGB). These were chosen because they are robust algorithms and well–established in clinical predictive modelling.[45] In perinatal medicine, preterm adverse outcomes frequently exhibit concomitance; for example, low Apgar scores are often associated with conditions such as hypoxaemic ischaemic encephalopathy and neonatal sepsis. Consequently, we opted against the development of a multiclass predictive model designed to identify each individual adverse outcome.

Each algorithm was trained using the transformed values of the seven FHR features. 10-fold cross validation was used, with each fold balanced for outcome, gestational age, trace duration and fetal sex. The optimum hyperparameters for each algorithm were identified using Bayesian optimisation. The average receiver-operator characteristic area under the curve (AUC) for each model was then used to evaluate the model’s overall performance. We ranked each model by AUC and compared the median AUC between each model with a Kruskal-Wallis one-way ANOVA and performed a pair-wise Mann-Whitney U test with a significance threshold of 0.01. The best-performing model was then selected as the final model for further analysis.

The model was then evaluated on the internal validation dataset using the AUC, sensitivity (of everyone classified as belonging to the preterm adverse outcome group, how many did the model correctly identify), specificity (how many normal FHR traces were correctly identified as such), F1 score (the harmonic mean of the proportion of true positives among the identified positives and sensitivity) and Cohen’s Kappa (the level of agreement between the predictive model and the known outcome). We determined that for a predictive model to demonstrate significant potential benefit, the average AUC must exceed 0.70 (in keeping with similar studies from the intrapartum period).[37, 46] An AUC of <0.6 would suggest poor discrimination, while 0.6–0.69 would be fair, 0.70–0.79 would be good and >0.8 would be excellent in keeping with similar publications.[47] We evaluated the AUC across all gestational ages and for each gestational age between 27^+0^ and 36^+0^ weeks.

Decision curve analysis was performed to compare the net benefit of the model against “treat-all” and “treat-none” strategies. In this context, a treat-all approach would be one in which pregnancies with suspected preterm adverse outcomes would be treated as such, e.g. delivered immediately. A treat-none strategy would comprise no further intervention in the pregnancy. Decision curve analysis evaluates how much “net benefit” the predictive model adds relative to these strategies. A robust predictive model should outperform the treat-all and treat-none strategies across a range of probability thresholds to identify pregnancies requiring treatment while avoiding unnecessary intervention in those that are unlikely to have an adverse outcome if untreated. An ineffective model would demonstrate either no net benefit or a lower net benefit relative to the treat-all strategy. We evaluated probability thresholds between 0.01 and 0.99 (1–99%). We then assessed the accuracy of the probability prediction (calibration degree) using the Brier score.

### Statistical Analysis

We adhere to TRIPOD guidelines for reporting.[48] Discrete variables are presented as numbers (with interquartile ranges) and percentages while continuous variables are listed as mean and 95% confidence intervals (95% CI). Categorical variables were compared using the Chi-square test while continuous variables were compared using the Mann-Whitney U test with a significant threshold of 0.05. Predictive models were compared using an ANOVA and pair-wise Mann-Whitney U test with a significance threshold of 0.01. P-values were estimated for each feature’s association with a high risk pregnancy using the Mann-Whitney U test and a significance threshold of 0.05. Confidence intervals were calculated using the bootstrap method. Effect sizes were analysed using Cohen’s D for parametric and Rank-biserial correlation for non-parametric variables. Analysis was performed using Python (version 3.9.17) with the Pandas (version 1.5.3), NumPy (version 1.23.5), Matplotlib (version 3.7.1) and SciPy (version 1.10.1) packages.

## Results

The study population comprised 8,881 FHR traces. 4,014 (45.2%) normal and 4,867 (54.8%) high-risk preterm adverse outcome traces were identified (Table 1). The median maternal age was 30 years (25–34), median parity was 1 (0–1) and median BMI was 23.5 (21.3–26.2). 4,335 (48.8%) traces were from male fetuses and 4,546 (51.2%) were from female fetuses. The distribution of outcomes did not differ significantly across gestational ages (p = 0.17). Significant differences were observed for all FHR features between the normal and preterm adverse outcome groups (Table 2). Median accelerations, episodes of high variation and short-term variation were significantly higher in the normal outcome group (p<0.001) while the basal heart rate, decelerations, episodes of low variation and most lost beats were significantly higher in the preterm adverse outcome group (p<0.001).

**Table 1:** Study population characteristics. Normal and preterm adverse outcome cohorts were developed to train machine learning algorithms to predict whether a fetal heart rate (FHR) trace belonged to either group. 4,014 FHR traces were identified for the normal cohort and 4,867 traces for the preterm adverse outcome cohort. Women in the preterm adverse outcome were on average ∼ 2 years older and had not yet experienced a viable pregnancy (effect size <0.01). They also demonstrated on average marginally higher body mass indices (BMI) however the effect size was medium (∼ 0.2). There was no significant difference in the fetal sex of the pregnancy between each group. The effect size for maternal age, viable and non-viable parity was small, indicating the absolute difference between the two groups was not substantial.

| Feature | Normal outcome | Preterm adverse outcome | $p$ | Effect size |
| --- | --- | --- | --- | --- |
| FHR traces | 4,014 | 4,867 | – | – |
| Maternal age | 29 (25–33) | 31 (26–35) | $<0.01$ | Small |
| Viable parity | 1 (0–1) | 0 (0–1) | $<0.01$ | Small |
| Non-viable parity | 0 (0–1) | 0 (0–1) | $<0.01$ | Small |
| BMI | 23.5 (21.2–25.9) | 24.9 (22.0–29.4) | $<0.01$ | Medium |
| Fetal sex | Male: 1,960<br>Female: 2,054 | Male: 2,375<br>Female: 2,492 | 0.99 | – |

**Table 2:** Comparison of fetal heart rate features between normal and preterm adverse outcome pregnancies. Fetal heart rate patterns (features) were extracted from all traces using an automated algorithm. Significant differences were observed between the normal and pre-term adverse outcome groups for each signal feature. Values shown are the median value for each feature with the interquartile range in brackets. The effect size was small for baseline heart rate and decelerations (≤ −0.10; ≥ 0.1), medium for most lost beats (≤ −0.3; ≥0.3) and large for accelerations, high variation minutes, low variation minutes and short-term variation (<-0.3; >0.3).

| Feature | Normal outcome | Preterm adverse outcome | *p* | Effect size |
| --- | --- | --- | --- | --- |
| Accelerations | 5 (3–8) | 3 (1–5) | <0.001 | -0.4 (Large) |
| Baseline heart rate | 138 (132–144) | 139 (132–145) | <0.001 | 0.0 (Small) |
| Decelerations | 0 (0–1) | 0 (0–1) | <0.001 | 0.1 (Small) |
| High variation (minutes) | 7 (3–14) | 2 (0–7) | <0.001 | -0.4 (Large) |
| Low variation (minutes) | 0 (0–1) | 4 (0–22) | <0.001 | 0.4 (Large) |
| Most lost beats | 8 (6–11) | 11 (8–18) | <0.001 | 0.2 (Medium) |
| Short-term variation | 9 (8–11) | 7 (5–9) | <0.001 | -0.5 (Large) |

The data were then split 80% into model training and 20% internal validation datasets, balanced for outcome, trace duration, gestational age and fetal sex. We trained the algorithms to predict the preterm adverse outcome group using the seven fetal heart rate features and 10-fold cross-validation with each fold balanced for outcome, trace duration, gestational age and fetal sex. We then compared the performance of each predictive model using the receiver-operator area under the curve (AUC) (Figure 2, Supplementary Table 2). The random forest and XGBoost algorithms demonstrated the best performance with a mean AUC of 0.88 (95% CI 0.87-0.88) and 0.87 (95% CI 0.86-0.87, p<0.001).

**Fig. 1:**
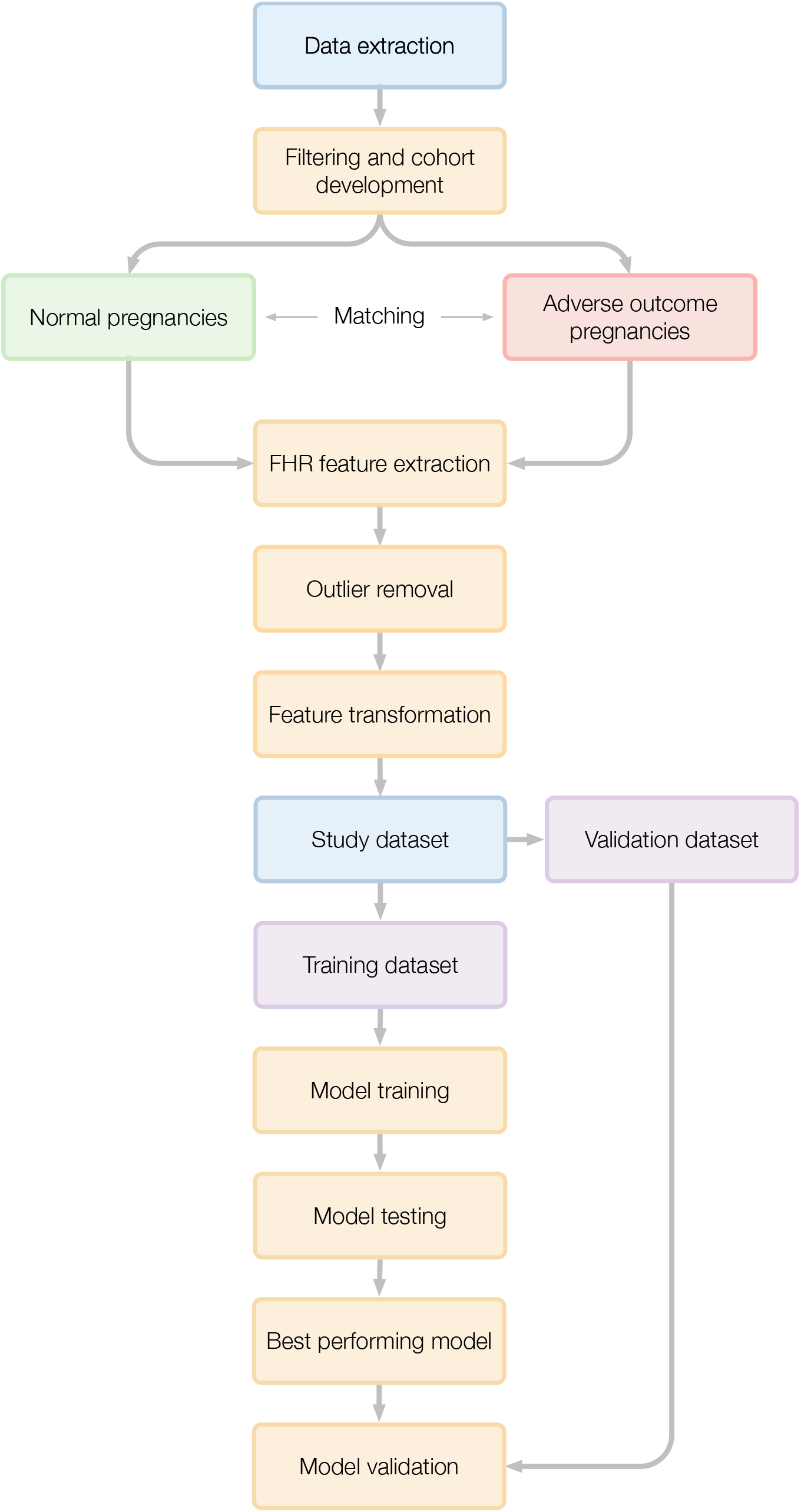
Data flow for the development of a study dataset and predictive model to identify pre-term adverse outcome pregnancies using the antepartum fetal heart rate.

**Fig. 2:**
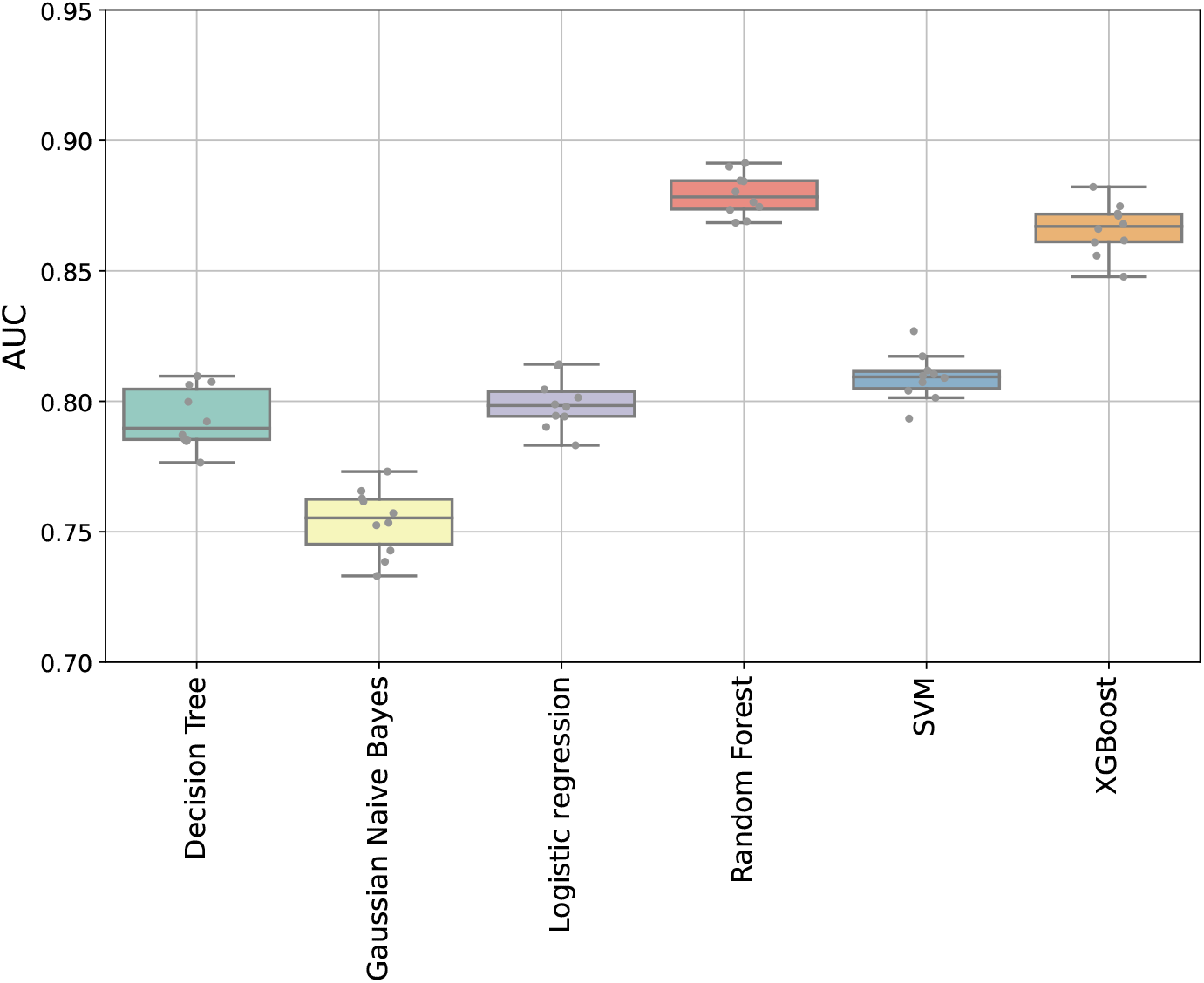
Comparison of the area under the curve (AUC) across six machine learning algorithms. Each algorithm was trained on the model training dataset with 10-fold cross validation. The best performing model was the random forest algorithm (AUC 0.88, IQR 0.87–0.88). AUC = receiver-operator area under the curve. See Supplementary Table 2 for values.

The relative importance of each FHR feature in the random forest model was subsequently evaluated. In a random forest model, the importance of each feature is determined by measuring how much a particular feature improves the model’s performance, averaged across all the trees within the forest. Importance varied considerably across the assessed features. The most salient predictor was short-term variation which accounted for 27.1% of the model’s predictive capacity. This was followed by baseline heart rate and episodes of high variation contributing 16.4% and 13.8% to the model’s accuracy, respectively. Features such as accelerations and episodes of low variation also held moderate predictive value, with importances of 12.0% and 11.2%, correspondingly. Conversely, most lost beats and decelerations had diminished relative importance, contributing merely 5.4% and 2.4% to the model’s overall predictive performance (Supplementary Figure 1).

The predictive performance of the random forest model was then evaluated for each of the individual outcomes contributing to a classification of preterm adverse outcome. The majority of outcomes exceeded an AUC of 0.80. The median AUC across all individual outcomes was 0.85 (IQR 0.81–0.89) demonstrating robust performance (Supplementary Table 3). The highest AUC was for hypoxic ischaemic encephalopathy (AUC 0.99, IQR 0.70–0.99, n=7) while the lowest was for a special care admission exceeding one week (AUC 0.77, IQR 0.73–0.80, n=161).

We then evaluated the model’s performance on the internal validation dataset. The model performed well with an AUC of 0.88 (95% CI 0.86–0.90, Figure 3) and demonstrated a high degree of calibration (Brier score 0.14, Supplementary Figure 2). Decision curve analysis was used to assess the net benefit of the model across a range of probability thresholds (0.01–0.99) compared to treat-all and treat-none strategies. The net benefit of the model exceeded the treat-all strategy for all probability thresholds above 0.11 (Supplementary Figure 3) and exceeded the treat-none strategy for all probability thresholds.

**Fig. 3:**
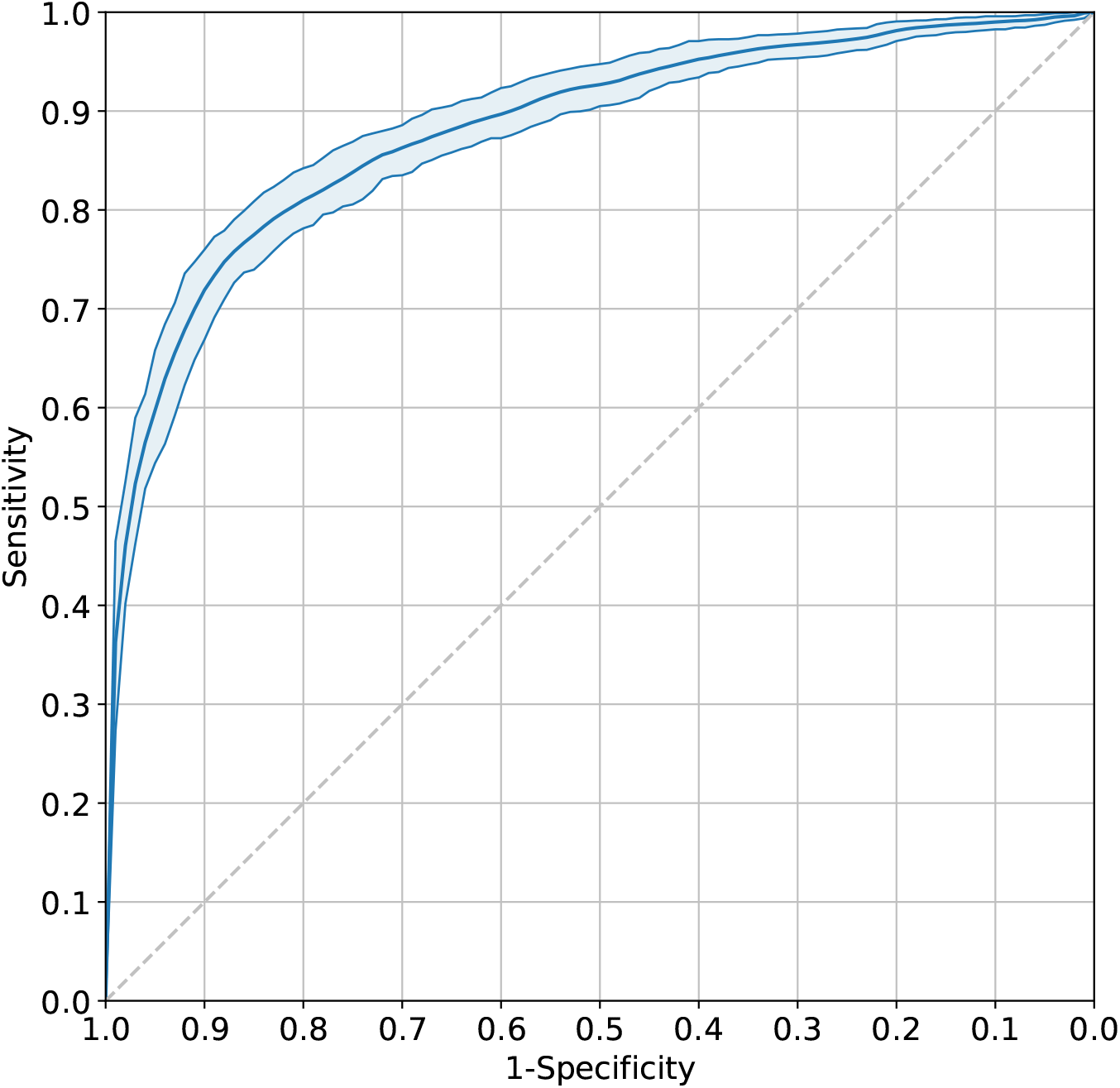
Receiver operating characteristic (ROC) curve for the prediction of an adverse outcome in a pre-term fetus on the validation dataset. The area under the curve (AUC) for the random forest classifier was 0.88 (95% CI 0.86–0.90), demonstrating an ‘excellent’ degree of performance.

Three probability thresholds for a classification of normal or preterm adverse outcome were evaluated: the Youden index (the position on the ROC curve whereby sensitivity and specificity are maximal) and the thresholds corresponding to 95% sensitivity and 95% specificity. For each threshold, we evaluated the sensitivity, specificity, F1 score and Cohen’s Kappa (Table 3). The sensitivity and specificity using the Youden index threshold (threshold = 59.6%, 95% CI 55.6–62.9%) was 76.2% (95% CI 72.6–80.5%) and 87.5% (95% CI 83.3–91.0). The specificity for the 95% sensitivity threshold (29.3%, 95% CI 28.9–31.6%) was 41.4% (95% CI 33.9–49.2%). The sensitivity for the 95% specificity threshold (73.6%, 95% CI 70.1–77.0%) was 59.3% (95% CI 54.0–65.6). The F1 scores for the three thresholds were 81.7 (95% CI 79.6–83.9), 78.1 (95% CI 75.4–80.7) and 72.5 (95% CI 68.3–77.4). Cohen’s Kappa was 62.8 (95% CI 59.6–66.4), 38.1 (95% CI 30.3–46.4) and 52.3 (95% CI 46.6–58.6).

**Table 3:** Evaluation of the random forest model using the validation dataset. The performance of the predictive model was evaluated using three different probability thresholds. Cases above the threshold were classified as preterm adverse outcome pregnancies, those below were designated as normal. The Youden threshold is the probability threshold at which the sensitivity and specificity are maximal. Sensitivity (true positive rate) measures the proportion of actual preterm adverse pregnancy outcomes that are correctly identified by the model. Specificity (true negative rate) quantifies the proportion of actual normal outcome pregnancies accurately classified by the model. The F1 score is the harmonic mean of precision (the proportion of true positives among the identified positives) and sensitivity, providing a balanced measure of a model’s performance. Cohen’s Kappa measures the degree to which the classifications made by the algorithm agree with the true classifications, accounting for the agreement that would be expected by random chance. Values in brackets denote the 95% confidence intervals.

|  | Threshold (%) | Sensitivity (%) | Specificity (%) | F1 score (%) | Cohen's Kappa |
| --- | --- | --- | --- | --- | --- |
| Youden | 59.6<br>(55.6–62.9) | 76.2<br>(72.6–80.5) | 87.5<br>(83.3–91.0) | 81.7<br>(79.6–83.9) | 62.8<br>(59.6–66.4) |
| 95% Sensitivity | 29.3<br>(28.9–31.6) | – | 41.4<br>(33.9–49.2) | 78.1<br>(75.4–80.7) | 38.1<br>(30.3–46.4) |
| 95% Specificity | 73.6<br>(70.1–77.0) | 59.3<br>(54.0–65.6) | – | 72.5<br>(68.3–77.4) | 52.3<br>(46.6–58.6) |

We then assessed the performance of the model for each gestational age interval (Supplementary Table 4 & Supplementary Figure 4). The median AUC exceeded 0.90 between 27^+0^ (AUC 0.93, 95% CI 0.86-0.98) and 31^+6^ weeks (AUC 0.93, 95% CI 0.89–0.97) and exceeded 0.80 for all subsequent weeks. The highest AUC observed was 0.93 (95% CI 0.89–0.97) at 31^+0^–31^+6^ weeks while the lowest was at 36^+0^–36^+6^ weeks (0.81, 95% CI 0.77–0.85).

## Discussion

We have shown machine learning algorithms can contribute substantially towards identifying high-risk pre-term pregnancies using antepartum FHR patterns. We identified a cohort of high-risk pre-term pregnancies and used a clinically-validated algorithm to extract seven physiologically-validated fetal heart rate features that were independent from the pitfalls of subjective assessment. We then applied machine learning algorithms to develop a high-fidelity predictive model capable of discriminating across a range of gestational ages. The model performed well when evaluated across a range of metrics on the validation dataset, including high sensitivity, specificity, F1 score and Cohen’s kappa. Decision curve analysis also demonstrated the model significantly outperformed both treat-all and treat-none strategies.

To our knowledge, this is the first study of its kind. This study employs a fully-automated machine-learning approach, independent from subjective clinical evaluation. We also utilised discrete, objective clinical outcomes to define a high-risk pre-term cohort. Previous studies have either used FHR patterns identified by visual interpretation or trained algorithms to predict outcomes based on clinical impressions (for example a “non-reassuring trace”), both of which are susceptible to subjectivity and bias. This represents an important advancement in the application of machine learning to clinical care of the pregnancy. FHR monitoring is one of the few yet affordable technologies available for immediate and real-time evaluation of fetal physiology and wellbeing. Human clinical experts have frequently proven limited at this task. These signals are complex and difficult to interpret visually, resulting in poor inter- and intra-rater reliability and high false positive rates. This has been associated with avoidable caesarean sections (and consequent preterm morbidity), as well as adverse fetal outcomes from a failure to intervene. These results indicate machine learning algorithms possess substantial promise in outperforming clinical experts and existing systems at this task. Early detection of high-risk pre-term pregnancies is critical. Exposure to labour would place these pregnancies at a substantially increased risk of an adverse outcome or death. Therefore, a high-fidelity system decoupled from the difficulties and inherent biases associated with visual interpretation of these signals would be of significant benefit.

The relationships we identified between each FHR pattern and the prediction of a high-risk pre-term pregnancy are supported by previous studies. FHR accelerations are an indicator of neurological health, concomitant with a healthy response to transient umbilical cord compression and fetal movements and suggest an absence of hypoxia.[49] Episodes of high variation are analogous to episodes of active sleep-/wakefulness. Cycling between episodes of active sleep is a hallmark of normal neurological development, the absence of which has been associated with acidaemia, hypoxia and low Apgar scores.[49, 50] Low STV values have been associated with an increased risk of fetal acidaemia.[51, 52] Below normal basal fetal heart rates (fetal bradycardia) are associated with an increased risk of adverse outcomes, including hypoxia, sustained umbilical cord compression, hypoxia, cardiac anomalies and maternal hypotension.[53, 54] Decelerations and their magnitude are associated with an increased risk of an adverse pregnancy outcome. Large magnitude decelerations are known to occur in acute fetal hypoxia and acidosis.[55] Episodes of low variation are analogous to quiet/deep sleep. Prolonged episodes in the absence of high variation episodes suggest a potential deficiency in normal neurological development.[44] We also demonstrated that a maximally-discriminative model should incorporate all of these patterns. This is in contrast to some current clinical guidelines which only employ a subset of features.[49, 56] Frequently, one or more of these patterns are absent from a trace (e.g. accelerations or decelerations) yet do not necessarily convey an increased risk of adverse outcome.[49] In this case, a multivariate approach incorporating other such patterns is required. Historically, a failure to recognise this has resulted in unnecessary deliveries.[57, 58]

The predictive performance of our model mildly declined after 35^+0^ gestational weeks. These changes potentially reflect the current understanding of the physiological development of FHR patterns. Towards term, decelerations occur more frequently in normal pregnancies, subsets of which are an indicator of normal physiological responses to transient cord compression for example.[59] As the normal neurological system of the fetus develops, cycling between quiet and active sleep also occurs more frequently with the mean duration of an episode of low variation increasing.[60– 62] The average short-term variability also increases with gestational age.[63]

Most studies developing machine learning models analysing the FHR have focussed on the intrapartum period.[37, 64] Some studies attempted to develop predictive models using fetal heart rate patterns identified by human visual inspection of the FHR trace, limiting the reproducibility of these models due to the inherent problems with human evaluation.[46] Other studies have either utilised significantly smaller datasets, did not use established clinical outcomes or were developed using a restricted subset of pregnancies.[65–67] Some were designed to assign the fetal heart rate into international classifications already known to suffer from poor performance. One study incorporated only the last 30 minutes of a FHR trace.[37] This substantially restricts generalisability and introduces bias, as the length of a trace prior to acquisition is frequently unknowable and longer traces are generally performed because evaluation of the trace during acquisition has resulted in continued recording.

While deep neural network architectures are a clear consideration for analysing such data and future plans include these studies, there are several important benefits to this current approach. The FHR patterns in our model are physiologically-driven and their importance in the model easily interpretable, enabling simplified interrogation of results and the potential to advance our understanding of these patterns. These results will also serve as an important first benchmark for future studies. A simple, understandable algorithm relying on low-cost and accessible technology also makes these algorithms more widely implementable and available to more regions. FHR monitoring is one of only a few technologies available that offers real-time appraisal of fetal physiology. Other modalities are either costly, time consuming, require extensive training or are inaccessible to many. While this technology is more affordable in low-resource regions, the requirement for adequate clinical training and expertise remains substantial. In such a setting, robust and effective machine learning algorithms could serve an important purpose.

## Conclusions

There is overwhelming consensus that electronic fetal heart rate monitoring is an important and irreplaceable investigation in the assessment of fetal wellbeing. Clinical experts and current technologies are considerably limited in interrogating this information, often resulting in avoidable adverse outcomes. We have developed a predictive model demonstrating machine learning possesses significant capability in accurately detecting high-risk preterm pregnancies. This bespoke model may enable earlier and more accurate diagnosis and facilitate better management of these pregnancies.

## Data Availability

The authors acknowledge the importance of data transparency and the potential value of data sharing in advancing scientific research. However, due to the identifiable and sensitive nature of the data used in this study, which includes detailed fetal heart rate
traces potentially linked to individual patient outcomes, we are unable to make the dataset publicly available. The data contains protected health information and is subject to strict confidentiality constraints to safeguard the privacy of individuals. Consequently, the ethical and legal restrictions prevent the sharing of the dataset.

## References

[1] Cohen, W. R. et al. Accuracy and reliability of fetal heart rate monitoring using maternal abdominal surface electrodes. Acta obstetricia et gynecologica Scandinavica 91, 1306–1313 (2012).

[2] Giussani, D. A., Spencer, J. A., Moore, P. J., Bennet, L. & Hanson, M. A. Afferent and efferent components of the cardiovascular reflex responses to acute hypoxia in term fetal sheep. The Journal of physiology 461, 431–449 (1993). URL https://pubmed.ncbi.nlm.nih.gov/8350271https://www.ncbi.nlm.nih.gov/pmc/articles/PMC1175265/.

[3] Benarroch, E. E. Control of the cardiovascular and respiratory systems during sleep. Autonomic Neuroscience 218, 54–63 (2019).

[4] Baan, J. J., Boekkooi, P. F., Teitel, D. F. & Rudolph, A. M. Heart rate fall during acute hypoxemia: a measure of chemoreceptor response in fetal sheep. J Dev Physiol 19, 105–11 (1993).

[5] Horne, R. S. Autonomic cardiorespiratory physiology and arousal of the fetus and infant. University of Adelaide Press (2018).

[6] Preboth, M. Acog guidelines on antepartum fetal surveillance. American family physician 62, 1184–1188 (2000).

[7] Ahn, M. O., Phelan, J. P., Smith, C. V., Jacobs, N. & Rutherford, S. E. Antepartum fetal surveillance in the patient with decreased fetal movement. American journal of obstetrics and gynecology 157, 860–864 (1987).

[8] Davis Jones, G., Albert, B., Cooke, W. & Vatish, M. A performance evaluation of computerised antepartum fetal heart rate monitoring: The dawes-redman algorithm at term. medRxiv 2024–02 (2024).

[9] Hammacher, K. New method for the selective registration of the fetal heart beat. Geburtshilfe und Frauenheilkunde 22, 1542 (1962).

[10] Gagnon, R., Campbell, M. K. & Hunse, C. A comparison between visual and computer analysis of antepartum fetal heart rate tracings. American journal of obstetrics and gynecology 168, 842–847 (1993).

[11] Todros, T., Preve, C., Plazzotta, C., Biolcati, M. & Lombardo, P. Fetal heart rate tracings: observers versus computer assessment. European Journal of Obstetrics & Gynecology and Reproductive Biology 68, 83–86 (1996).

[12] Beaulieu, M. et al. The reproducibility of intrapartum cardiotocogram assessments. Canadian Medical Association Journal 127, 214 (1982).

[13] Borgatta, L., Shrout, P. E. & Divon, M. Y. Reliability and reproducibility of nonstress test readings. American journal of obstetrics and gynecology 159, 554– 558 (1988).

[14] Chandraharan, E. Handbook of CTG interpretation: from patterns to physiology (Cambridge University Press, 2017).

[15] Bernardes, J., Costa-Pereira, A., Ayres-de Campos, D., Geijn, H. & Pereira-Leite, L. Evaluation of interobserver agreement of cardiotocograms. International Journal of Gynecology & Obstetrics 57, 33–37 (1997). URL https://obgyn.onlinelibrary.wiley.com/doi/abs/10.1016/S0020-7292%2897%2902846-4.

[16] Iams, J. D. Assessment and care of the fetus: Physiological, clinical, and medicolegal principles. JAMA 264, 2451–2451 (1990).

[17] Freeman, R. K., Anderson, G. & Dorchester, W. A prospective multi-institutional study of antepartum fetal heart rate monitoring. i. risk of perinatal mortality and morbidity according to antepartum fetal heart rate test results. Am J Obstet Gynecol 143, 771–7 (1982).

[18] Bobitt, J. R. Abnormal antepartum fetal heart rate tracings, failure to intervene, and fetal death: review of five cases reveals potential pitfalls of antepartum monitoring programs. Am J Obstet Gynecol 133, 415–21 (1979).

[19] Williams, B. & Arulkumaran, S. Cardiotocography and medicolegal issues. Best Practice & Research Clinical Obstetrics & Gynaecology 18, 457–466 (2004).

[20] Sinai Talaulikar, V. & Arulkumaran, S. Medico-legal issues with ctg interpretation. Current Women’s Health Reviews 9, 145–157 (2013).

[21] Ayres-de-Campos, D., Bernardes, J., Costa-Pereira, A. & Pereira-Leite, L. Inconsistencies in classification by experts of cardiotocograms and subsequent clinical decision. BJOG: An International Journal of Obstetrics & Gynaecology 106, 1307–1310 (1999).

[22] Flynn, A. M. & Kelly, J. Evaluation of fetal wellbeing by antepartum fetal heart monitoring. British Medical Journal 1, 936–939 (1977). URL https://www.bmj.com/content/bmj/1/6066/936.full.pdf.

[23] Lyons, E., Bylsma-Howell, M., Shamsi, S. & Towell, M. A scoring system for non-stressed antepartum fetal heart rate monitoring. American Journal of Obstetrics and Gynecology 133, 242–246 (1979).

[24] Pearson, J. & Weaver, J. B. A six-point scoring system for antenatal cardiotocographs. BJOG: An International Journal of Obstetrics & Gynaecology 85, 321–327 (1978).

[25] Flynn, A. M., Kelly, J., Matthews, K., O’CONOR, M. & Viegas, O. Predictive value of, and observer variability in, several ways of reporting antepartum cardiotocographs. BJOG: An International Journal of Obstetrics & Gynaecology 89, 434–440 (1982).

[26] Valderrama, C. E., Ketabi, N., Marzbanrad, F., Rohloff, P. & Clifford, G. D. A review of fetal cardiac monitoring, with a focus on low-and middle-income countries. Physiological measurement 41, 11TR01 (2020).

[27] Organization, W. H. WHO recommendations on antenatal care for a positive pregnancy experience (World Health Organization, 2016).

[28] Das, M. K. et al. Clinical validation of mobile cardiotocograph device for intrapartum and antepartum monitoring compared to standard cardiotocograph: An inter-rater agreement study. Journal of Family & Reproductive Health 13, 109 (2019).

[29] Caly, H. et al. Machine learning analysis of pregnancy data enables early identification of a subpopulation of newborns with asd. Scientific reports 11, 1–14 (2021).

[30] Sufriyana, H. et al. Comparison of multivariable logistic regression and other machine learning algorithms for prognostic prediction studies in pregnancy care: systematic review and meta-analysis. JMIR medical informatics 8, e16503 (2020).

[31] Davidson, L. & Boland, M. R. Towards deep phenotyping pregnancy: a systematic review on artificial intelligence and machine learning methods to improve pregnancy outcomes. Briefings in bioinformatics 22, bbaa369 (2021).

[32] Schmidt, L. J. et al. A machine-learning–based algorithm improves prediction of preeclampsia-associated adverse outcomes. American Journal of Obstetrics and Gynecology (2022).

[33] Arnaout, R. et al. An ensemble of neural networks provides expert-level prenatal detection of complex congenital heart disease. Nature medicine 27, 882–891 (2021).

[34] Bertini, A., Salas, R., Chabert, S., Sobrevia, L. & Pardo, F. Using machine learning to predict complications in pregnancy: A systematic review. Frontiers in bioengineering and biotechnology 9 (2021).

[35] Chudáček, V. et al. Assessment of features for automatic ctg analysis based on expert annotation. Annual International Conference of the IEEE Engineering in Medicine and Biology Society. IEEE Engineering in Medicine and Biology Society. Annual International Conference 2011, 6051–6054 (2011).

[36] Warrick, P. A., Hamilton, E. F., Kearney, R. E. & Precup, D. A machine learning approach to the detection of fetal hypoxia during labor and delivery. Ai Magazine 33, 79–79 (2012).

[37] Petrozziello, A., Redman, C. W., Papageorghiou, A. T., Jordanov, I. & Georgieva, A. Multimodal convolutional neural networks to detect fetal compromise during labor and delivery. IEEE Access 7, 112026–112036 (2019).

[38] Morton, V. H. & Morris, R. K. Overview of the saving babies lives care bundle version 2. Obstetrics, Gynaecology & Reproductive Medicine 30, 298–300 (2020).

[39] Liu, L. et al. Global, regional, and national causes of under-5 mortality in 2000–15: an updated systematic analysis with implications for the sustainable development goals. The Lancet 388, 3027–3035 (2016).

[40] Blencowe, H. et al. National, regional, and worldwide estimates of preterm birth rates in the year 2010 with time trends since 1990 for selected countries: a systematic analysis and implications. The lancet 379, 2162–2172 (2012).

[41] Yudkin, P. L., Aboualfa, M., Eyre, J. A., Redman, C. W. & Wilkinson, A. R. New birthweight and head circumference centiles for gestational ages 24 to 42 weeks. Early Hum Dev 15, 45–52 (1987).

[42] Fetus, A. A. o. P. C. o. et al. The apgar score. Pediatrics 136, 819–822 (2015).

[43] Wu, P. et al. Developing and evaluating mappings of icd-10 and icd-10-cm codes to phecodes. bioRxiv 462077 (2018). URL https://www.biorxiv.org/content/biorxiv/early/2018/11/12/462077.full.pdf.

[44] Jones, G. D., Cooke, W. R., Vatish, M. & Redman, C. W. Computerized analysis of antepartum cardiotocography: A review. Maternal-Fetal Medicine 4, 130–140 (2022).

[45] Rosenfeld, A. et al. Development and validation of a risk prediction model to diagnose barrett’s oesophagus (mark-be): a case-control machine learning approach. The Lancet Digital Health 2, e37–e48 (2020).

[46] Ogasawara, J. et al. Deep neural network-based classification of cardiotocograms outperformed conventional algorithms. Scientific reports 11, 1–9 (2021).

[47] Polo, T. C. F. & Miot, H. A. Use of roc curves in clinical and experimental studies. J Vasc Bras 19, e20200186 (2020). 1677-7301 Polo, Tatiana Cristina Figueira Orcid: 0000-0001-9496-1053 Miot, Hélio Amante Orcid: 0000-0002-2596-9294 Editorial Brazil 2021/07/03 J Vasc Bras. 2020 Dec 11;19:e20200186. doi: 10.1590/1677-5449.200186.

[48] Collins, G. S., Reitsma, J. B., Altman, D. G. & Moons, K. G. Transparent reporting of a multivariable prediction model for individual prognosis or diagnosis (tripod) the tripod statement. Circulation 131, 211–219 (2015).

[49] Ayres-de-Campos, D., Spong, C. Y. & Chandraharan, E. Figo consensus guidelines on intrapartum fetal monitoring: Cardiotocography. International Journal of Gynecology & Obstetrics 131, 13–24 (2015).

[50] Pereira, S., Lau, K., Modestini, C., Wertheim, D. & Chandraharan, E. Absence of fetal heart rate cycling on the intrapartum cardiotocograph (ctg) is associated with intrapartum pyrexia and lower apgar scores. The Journal of Maternal-Fetal & Neonatal Medicine 1–6 (2021).

[51] Anceschi, M. M. et al. Validity of short term variation (stv) in detection of fetal acidemia. Journal of Perinatal Medicine 31 (2003).

[52] Henson, G., Dawes, G. & Redman, C. Antenatal fetal heart-rate variability in relation to fetal acid-base status at caesarean section. BJOG: An International Journal of Obstetrics & Gynaecology 90, 516–521 (1983).

[53] Hon, E. H. Observations on “pathologic” fetal bradycardia. American Journal of Obstetrics and Gynecology 77, 1084–1099 (1959).

[54] Jaeggi, E. T. & Friedberg, M. K. Diagnosis and management of fetal bradyarrhythmias. Pacing and clinical electrophysiology 31, S50–S53 (2008).

[55] Cahill, A. G., Roehl, K. A., Odibo, A. O. & Macones, G. A. Association and prediction of neonatal acidemia. American journal of obstetrics and gynecology 207, 206. e1–206. e8 (2012).

[56] Santo, S. et al. Agreement and accuracy using the figo, acog and nice cardiotocography interpretation guidelines. Acta Obstetricia et Gynecologica Scandinavica 96, 166–175 (2017). URL https://obgyn.onlinelibrary.wiley.com/doi/abs/10.1111/aogs.13064.

[57] Holzmann, M., Wretler, S. & Nordström, L. Absence of accelerations during labor is of little value in interpreting fetal heart rate patterns. Acta Obstetricia et Gynecologica Scandinavica 95, 1097–1103 (2016).

[58] Grivell, R. M., Alfirevic, Z., Gyte, G. M. L. & Devane, D. Antenatal cardiotocography for fetal assessment. Cochrane Database of Systematic Reviews (2015). URL 10.1002/14651858.CD007863.pub4.

[59] Dawes, G. et al. Large fetal heart rate decelerations at term associated with changes in fetal heart rate variation. American journal of obstetrics and gynecology 168, 105–111 (1993).

[60] Parmelee, A. & Stern, E. Development of states in infants. Sleep and the maturing nervous system 199–228 (1972).

[61] Saper, C. B., Scammell, T. E. & Lu, J. Hypothalamic regulation of sleep and circadian rhythms. Nature 437, 1257–1263 (2005).

[62] Sterman, M. & Hoppenbrouwers, T. The development of sleep-waking and rest-activity patterns from fetus to adult in man. Academic Press New York 203–227 (1971).

[63] Serra, V., Bellver, J., Moulden, M. & Redman, C. Computerized analysis of normal fetal heart rate pattern throughout gestation. Ultrasound in Obstetrics and Gynecology 34, 74–79 (2009).

[64] Ito, A. et al. Optimal duration of cardiotocography assessment using the ipreface score to predict fetal acidemia. Scientific Reports 12, 13064 (2022). URL 10.1038/s41598-022-17364-z.

[65] Zeng, R., Lu, Y., Long, S., Wang, C. & Bai, J. Cardiotocography signal abnormality classification using time-frequency features and ensemble cost-sensitive svm classifier. Computers in Biology and Medicine 130, 104218 (2021).

[66] Ayres-de Campos, D., Costa-Santos, C., Bernardes, J. & Group, S. M. V. S. Prediction of neonatal state by computer analysis of fetal heart rate tracings: the antepartum arm of the sisporto® multicentre validation study. European Journal of Obstetrics & Gynecology and Reproductive Biology 118, 52–60 (2005).

[67] Signorini, M. G., Pini, N., Malovini, A., Bellazzi, R. & Magenes, G. Integrating machine learning techniques and physiology based heart rate features for antepartum fetal monitoring. Computer methods and programs in biomedicine 185, 105015 (2020).

