## Supplementary Information for "Identifying high-risk pre-term pregnancies using the fetal heart rate and machine learning"

| Inclusion Criteria | Exclusion Criteria |
|---|---|
| Singleton pregnancy | Multiple pregnancy |
| Gestational age at delivery 37–41 weeks | Neonatal death within 3 months |
| Maternal age between 18–39 years | Premature spontaneous rupture of membranes |
| Maternal BMI ≤30 | Emergency caesarean section or breech presentation |
| Liveborn pregnancy | Unknown fetal sex at delivery |
| Duration of labour <24 hours | Arterial pH <7.13 and arterial base deficit >10.0 for babies delivered via caesarean section without labour |
| Apgar Score ≥4 at 1 minute and ≥7 at 5 minutes | arterial pH <7.05 and arterial base deficit >14.0 for babies who experienced labour (regardless of delivery method) |
| Umbilical arterial ± venous pH within normal range at delivery | Neonatal resuscitation required (any) |
| Cerebroplacental ratio >1.5 at 36 weeks | Special care admission following delivery |
| | Hypoxaemic ischaemic encephalopathy |
| | Neonatal cooling required |
| | Hypertensive disorders of pregnancy (e.g. pregnancy-induced hypertension, pre-eclampsia, etc.) or gestational diabetes |
| | Clinically-suspected or confirmed infection or sepsis |

Supplementary Table 1: Inclusion and exclusion criteria for the development of a cohort of ‘normal’ pregnancies.

| Algorithm | AUC (IQR) |
|---|---|
| Decision Tree | 0.79 (0.79-0.80) |
| Gaussian Naive Bayes | 0.76 (0.75-0.76) |
| Logistic regression | 0.80 (0.79-0.80) |
| Random Forest | 0.88 (0.87-0.88) |
| SVM | 0.81 (0.80-0.81) |
| XGBoost | 0.87 (0.86-0.87) |

Supplementary Table 2: Predictive performance of six machine learning algorithms trained to identify preterm adverse outcome FHR traces. Each algorithm was trained using 10-fold cross-validation. The median area under the curve (AUC) for each algorithm was then calculated using the AUC from each fold. The AUCs were compared using an ANOVA ($p<0.001$) and pairwise Mann-Whitney U test. The random forest algorithm performed best (AUC 0.88, IQR 0.87–0.88), outperforming the XGBoost algorithm (<0.001).

| Adverse outcome | Number of FHR traces | AUC (IQR) |
|---|---|---|
| Acidaemia | 215 | 0.85 (0.83-0.87) |
| Antepartum/Intrapartum stillbirth | 231 | 0.85 (0.82-0.89) |
| Asphyxia | 48 | 0.77 (0.69-0.85) |
| Birthweight ≤3rd centile | 3,030 | 0.86 (0.86-0.86) |
| Special care admission exceeding one week | 161 | 0.76 (0.73-0.80) |
| Hypoxaemic ischaemic encephalopathy | 7 | 0.99 (0.70-0.99) |
| Low Apgar score | 662 | 0.81 (0.79-0.82) |
| Neonatal sepsis | 39 | 0.93 (0.87-0.97) |
| Perinatal infections | 271 | 0.91 (0.89-0.91) |
| Respiratory conditions | 2,074 | 0.84 (0.83-0.86) |

**Supplementary Table 3: Evaluation of the predictive model by specific adverse outcome.** For each outcome within the pre-term adverse outcome classification, the AUC was evaluated, comparing the specific outcome against the normal cohort. The median AUC across all specific outcomes was 0.85 (IQR 0.81–0.89). The majority of outcomes exceeded an AUC of 0.80 ('excellent' performance). The highest AUC was for the hypoxaemic ischaemic encephalopathy outcome (AUC 0.99, IQR 0.70–0.99) while the lowest observed was for a special care admission exceeding one week (AUC 0.76, IQR 0.73–0.80).

| Gestational Age | Normal Outcomes | Preterm Adverse Outcomes | AUC (IQR) |
|---|---|---|---|
| $27^{+0}$–$27^{+6}$ | 34 | 39 | 0.93 (0.86-0.98) |
| $28^{+0}$–$28^{+6}$ | 35 | 81 | 0.92 (0.85-0.96) |
| $29^{+0}$–$29^{+6}$ | 49 | 83 | 0.91 (0.85-0.96) |
| $30^{+0}$–$30^{+6}$ | 58 | 77 | 0.91 (0.85-0.95) |
| $31^{+0}$–$31^{+6}$ | 63 | 90 | 0.93 (0.89-0.97) |
| $32^{+0}$–$32^{+6}$ | 74 | 101 | 0.88 (0.83-0.93) |
| $33^{+0}$–$33^{+6}$ | 83 | 115 | 0.87 (0.84-0.92) |
| $34^{+0}$–$34^{+6}$ | 105 | 103 | 0.87 (0.81-0.9) |
| $35^{+0}$–$35^{+6}$ | 126 | 120 | 0.86 (0.82-0.91) |
| $36^{+0}$–$36^{+6}$ | 176 | 163 | 0.81 (0.77-0.85) |

Supplementary Table 4: Evaluation of predictive model performance across gestational age intervals. Internal validation results for gestational age intervals, with AUC values ranging from 0.81 to 0.93. The highest AUC (0.93, 95% CI: 0.89-0.97) was observed at 31+0–31+6 weeks, and the lowest (0.81, 95% CI: 0.77-0.85) at 36+0–36+6 weeks. The dataset includes high-risk pre-term pregnancies with varying adverse outcomes, as detailed in Supplementary Table 2.

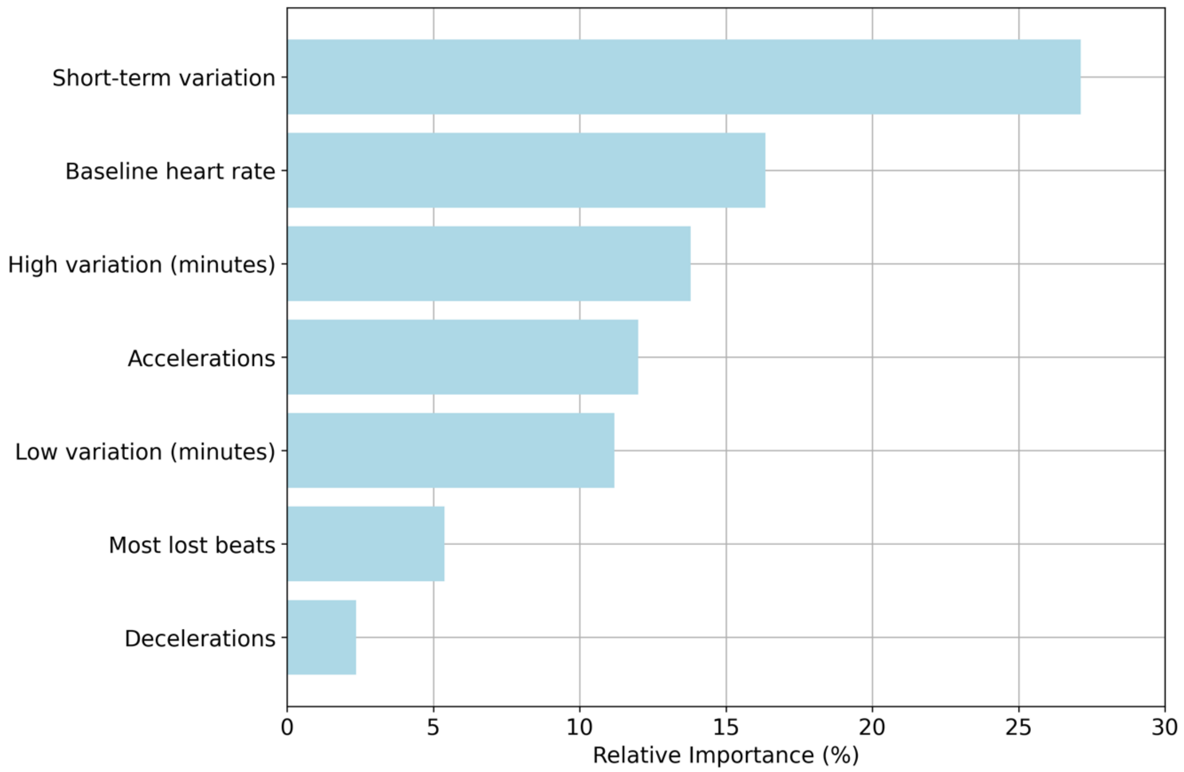


**Supplementary Figure 1: Relative importance of CTG features in predicting adverse pregnancy outcomes.** Relative feature importances are calculated as the average reduction in Gini impurity that each CTG feature contributes across all trees in the ensemble, normalized to sum to one. These importances serve as a quantitative measure to identify the most informative variables in predicting the outcome (normal or preterm adverse outcome). Short-term variation = 27.1%, baseline heart rate = 16.3%, high variation minutes = 13.8%, accelerations = 12.0%, low variation minutes = 11.2%, most lost beats = 5.4% and decelerations = 2.4%.

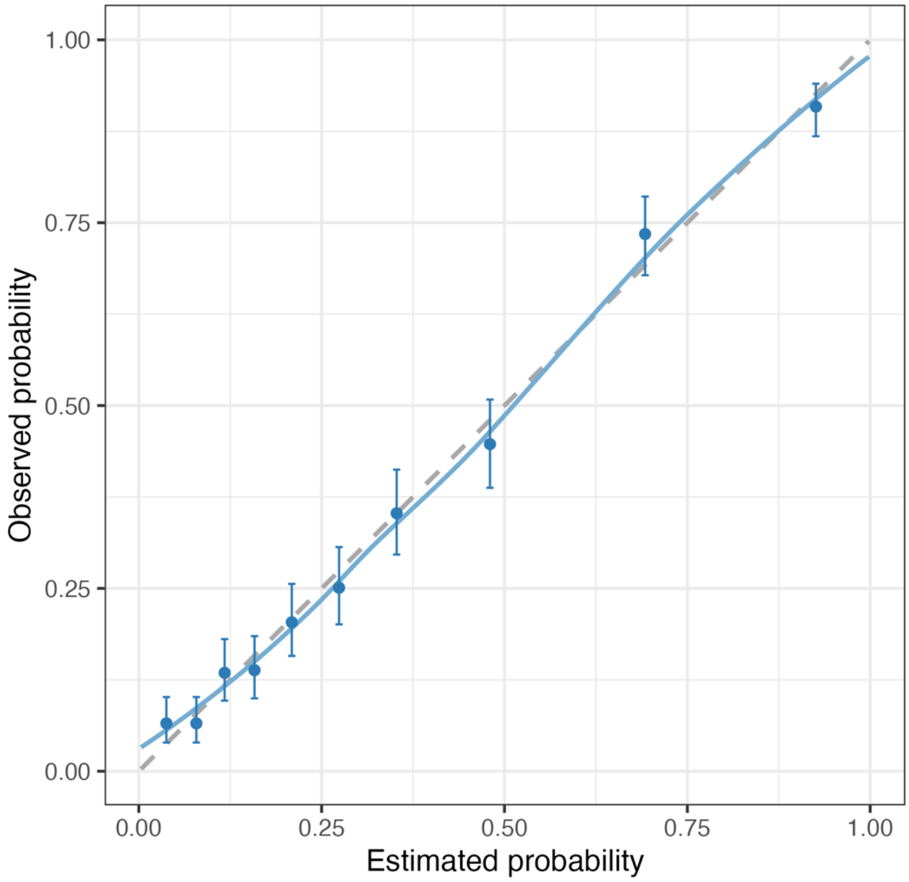


**Supplementary Figure 2: Calibration curve for the logistic regression model trained on a binarized dataset.** The model demonstrated a high degree of calibration using the Brier score: 0.15.

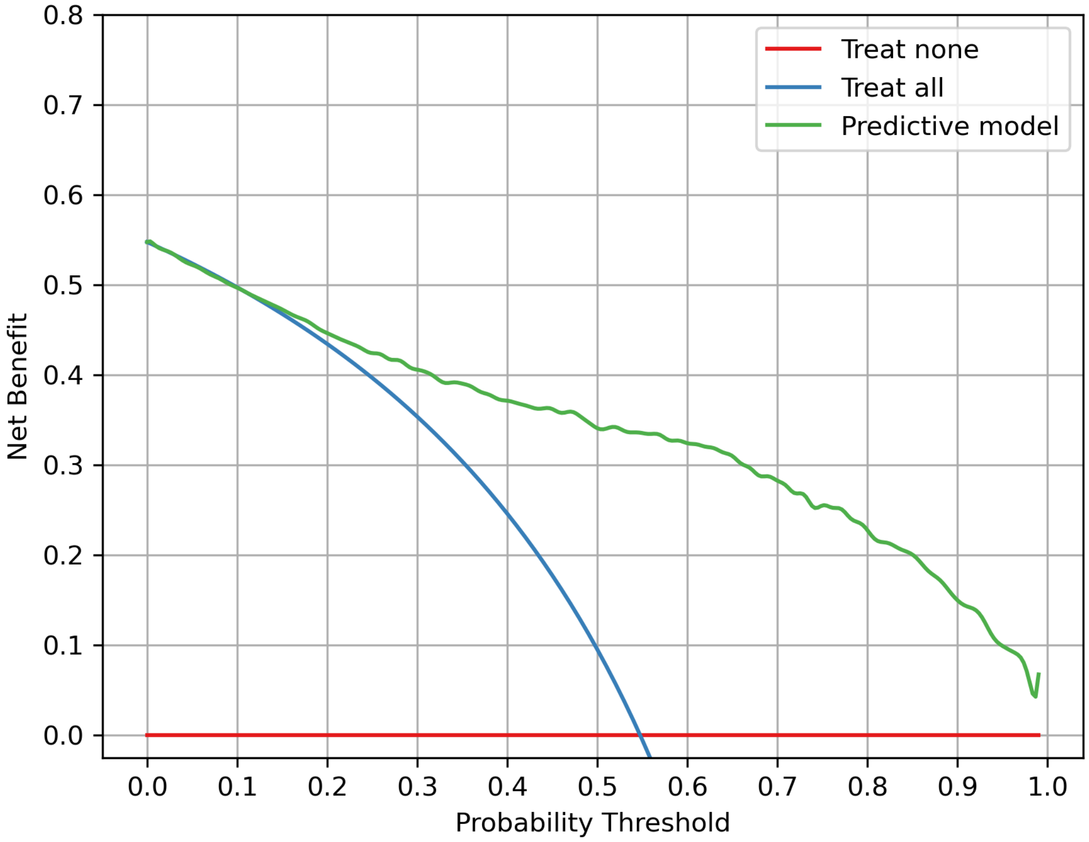


**Supplementary Figure 3: Decision curve analysis for a prediction of an adverse outcome in a pre-term pregnancy.** The net benefit of the predictive model (random forest) trained on a dataset of fetal heart rate features exceeded the treat none strategy across all probability thresholds and the treat all strategy for probabilities between 0.11–1.

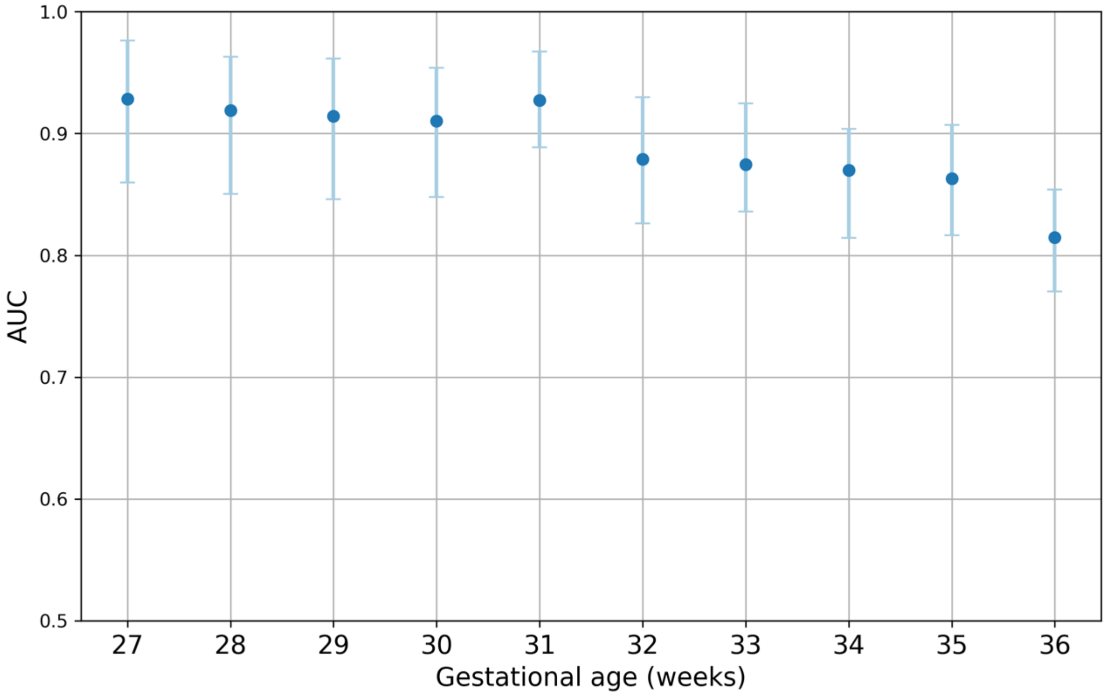


Supplementary Figure 4: Model performance with increasing gestational age. The random forest model was evaluated on the internal validation dataset at each gestational age interval (weeks). The model demonstrated excellent performance (AUC ≥0.80) across all weeks.
